# Direct intracranial EEG evidence for a local breakdown in normal sleep homeostasis at the human seizure onset zone

**DOI:** 10.1101/2025.11.04.25339396

**Authors:** Brinda Sevak, Dillon Scott, Edward Merricks, Colin Denis, Vaclav Kremen, Giulio Tononi, Catherine Schevon, Melanie Boly

## Abstract

**Background:** Animal studies have demonstrated that sleep is crucial for physiological synaptic pruning, and that this pruning is primarily regulated at a local, cellular level. We hypothesized that local alterations in sleep homeostatic regulation may constitute an interictal biomarker of the seizure onset zone (SOZ), and that sleep homeostasis abnormalities may also be present in areas beyond the SOZ, proportional to their secondary recruitment during seizures.

**Methods:** Overnight intracranial EEG (iEEG) recordings were obtained from sixteen well-characterized patients with focal epilepsy. The first and last hour of sleep were analyzed during one seizure-free night with well-organized sleep architecture, selected from each patient. We compared slow-wave activity (SWA; i.e., delta power, 1-4 Hz, a marker of synaptic strength) during wakefulness, the first and last hour of sleep, as well as SWA overnight decline in the SOZ, immediately neighboring regions, and regions located >2 cm away. Sleep-dependent changes in the slope of slow-waves and spike-waves were also considered. Finally, we examined whether sleep SWA and slow-wave slope abnormalities were predicted by ictal phase-locked high-gamma (PLHG), a proxy of ictal recruitment, both in the SOZ and in regions away from it.

**Results:** Consistent with our hypothesis, the SOZ displayed higher increases in SWA from wake to sleep than other brain regions. During sleep, SWA and slow-wave slope were higher in the SOZ compared to both immediately neighboring regions and regions further apart, with the difference being most pronounced during the last hour of sleep. Additionally, we observed a dampened overnight decline in SWA in the SOZ compared to distant cortical areas, suggesting an escape from physiological synaptic homeostasis. While the slopes of epileptic spike-waves were also higher in the SOZ compared to neighboring and distant areas, they did not significantly decline overnight in any area. Finally, as hypothesized, ictal PLHG (a marker of recruitment) predicted the extent of increases in sleep SWA, sleep slow-wave slope, and spike-wave slopes across all brain areas, with results especially consistent for the last hour of the night.

**Conclusion:** Our results provide direct intracranial EEG evidence for local alterations in sleep homeostasis in human epileptic brains, which are maximal at the SOZ, and proportional to local ictal recruitment across the whole brain. These results provide a solid mechanistic foundation for interventional studies aiming to enhance local sleep homeostasis in epileptic brains, through e.g. pharmacology or neuromodulation, to decrease local neuronal excitability and improve patient outcomes.

## Introduction

Epilepsy is a disabling neurological condition that affects over 1% of people worldwide.^1^ Among adults, the most common form is focal epilepsy, where seizures originate from a specific area in the brain. However, the diagnosis of focal epilepsy (FE) is often delayed. About 60% of patients with FE become seizure free with the use of anti-epileptic drugs (AEDs), however, more than 30-40% patients have drug resistant epilepsy (DRE).^1^ Resective surgery is the current choice of treatment for DRE patients, however, it does not always make the patient seizure free. A better understanding of the mechanisms behind increases in neuronal excitability leading to FE is crucial to the improvement of its diagnosis and treatment.

Currently, the cellular mechanisms leading to the occurrence of ongoing seizures in the human brain are not completely known ^2^. A potential candidate mechanism is the induction of long-term potentiation by both epileptic spikes and seizures, ^3^ as suggested by several animal studies ^4-8^. This finding is also in line with human slice work has also shown a 2.6-fold increase in synaptic density with increase in CREB expression (a marker of long-term potentiation) in layers 2/3 of epileptic cortex compared to neighboring cortex.^9-11^ In animal studies, epileptic activity has also been shown to affect memory formation via competition with normal plastic changes involved in information processing, potentially contributing to poor cognitive outcomes. ^3^ Likewise, human patients with epilepsy show a decline in cognitive performance along with disruption in memory formation which is proportional to the seizure burden created by their epileptic disease.^12^

Previous animal studies have validated slow-wave activity (SWA, i.e. delta power, 1-4 HZ) during sleep to be a reliable marker of synaptic strength.^13^ Previous work from our team using intracranial EEG in both humans and animals has also shown that most sleep slow waves are local,^14,15^ and homeostatic regulation of SWA during sleep (reflected by SWA overnight decline) is primarily regulated at a local level as well.^16,17^ More recently, electron microscopy studies in mice have provided a direct demonstration of the necessary role of sleep for a net decrease in synaptic strength over the course of the night (with up to 20% decrease in synaptic strength over the course of one night) both in cortical and subcortical areas (e.g. seen across the neocortex,^18^ hippocampus^19^ and cerebellum ^20^). A crucial role of sleep for whole-brain synaptic pruning was recently confirmed by an independent group in the Zebrafish.^21^ Importantly, synaptic pruning has also been shown to only occur consistently for the 80% smallest cortical synapses, and not for the 20% strongest ones.^18^ While such selective synaptic homeostasis plasticity rules may usually be adaptive by helping to preserve important memories in neurotypical brains, it may be prone to hijacking within hyperconnected epileptic networks, favoring the persistence of pathological hyperexcitability.^3^ Additional findings from animal studies suggest that while neuronal firing during the up-state of the slow-waves is necessary for synaptic pruning, bursting firing patterns may by themselves prevent downscaling.^22^ Such a paradoxical effect of bursting firing patterns on synaptic homeostasis may partly explain the negative association between frequent interictal discharges during and sleep overnight SWA decline previously observed in patients with both adult or pediatric epilepsy ^3,23,24^. Comparing sleep-dependent changes in SWA and spike-wave activity within the epileptic focus vs the rest of the cortex may thus provide valuable insights into mechanisms leading to persistent network excitability in human patients with FE.

Previous scalp EEG studies in humans have shown broad increases in SWA overnight and insufficient overnight decline, with maxima seemed to colocalize with the likely seizure foci on scalp EEG ^3,23,24^. However, such studies had insufficient resolution to compare sleep homeostasis markers at the seizure onset zone (SOZ) vs. neighboring cortical areas. Additionally, such scalp studies lacked the ability to directly link local SWA activity changes during sleep to validated intracranial markers of ictal recruitment such as local ictal phase-locked gamma activity (PLHG, ^25,26^). While our previous intracranial EEG work identified some local increases in sleep SWA in the SOZ compared to regions > 2cm from it during randomly sampled epochs of sleep,^27^ it did not provide a comprehensive analysis of differences in dynamical changes in SWA in terms of overnight decline. This is important because unlike pathological increases in SWA, physiological learning-dependent increases in SWA at the beginning of the night are expected to be accompanied by an overall increase in local overnight SWA decline in the same areas.^28^ The observation of a combination of locally increased SWA with dampened overnight SWA decline would provide strong arguments for an escape from sleep homeostasis being one of the potential contributing mechanism for persistent hyperexcitability in the seizure focus. Our previous intracranial EEG study also had not examined relationships between sleep SWA abnormalities and ictal PLHG patterns. The present study aims to resolves these issues by quantifying markers of sleep homeostasis using a gold-standard analysis of a whole-night of sleep SWA using direct intracranial recordings, also comparing overnight changes in the slope and amplitude of sleep slow-waves vs. epileptic spike-waves, and by directly testing for an association between local abnormalities in interictal markers of sleep homeostasis and local ictal recruitment as measured by PLHG during seizures. We believe that results obtained further consolidate the evidence for a strong association between local sleep homeostasis dysfunction and focal epileptogenic burden, which may pave new ways towards translational neuromodulation approaches to enhance sleep homeostasis locally at the SOZ in patients with FE, aiming to improve their clinical outcomes.

## Methods

### Human subjects

Our sample consisted of a total of sixteen patients with drug resistant focal epilepsy [mean age 29 ± standard deviation (SD) 11, eight females, seven males] undergoing invasive EEG study to help identify the seizure onset zone (SOZ) for resection. The SOZ were localized either in the left temporal (n = 5), left frontal operculum (n=1), right temporal (n = 7), right insula (n =1) or bilateral temporal (n = 1) regions. Clinical determinations of the SOZ were made by treating physicians and confirmed by a qualified epileptologist (C.A.S.) prior to analysis. Inclusion criteria was limited to patients with good localization of the SOZ in the presurgical evaluation as well as a night of sleep that was seizure-free and with well-defined sleep cycle architecture.

Research was conducted under the guidance of Columbia University Medical Center’s Institutional Review Board (IRB) and each patient provided informed consent prior to the implantation. The study involved two cohorts: Four patients implanted with electrodes (3.0-mm diameter) with 0.5-or 1-cm center-to-center spacing (Representative patient Fig. 1Ai); Twelve patients implanted with depth (2.3-mm length) electrodes (Ad-Tech, Racine, WI) (Representative patient Fig. 1Aii). In both the cases, EEG signals were acquired using a standard clinical video EEG system (XLTek, Natus Medical Inc., Oakville, ON, Canada) at 500 Hz per channel (0.5–125 Hz bandpass, 24-bit precision). The implant site was selected from presurgical EEG studies, and eloquent cortical sites such as Broca’s area were avoided.

**Figure 1.**
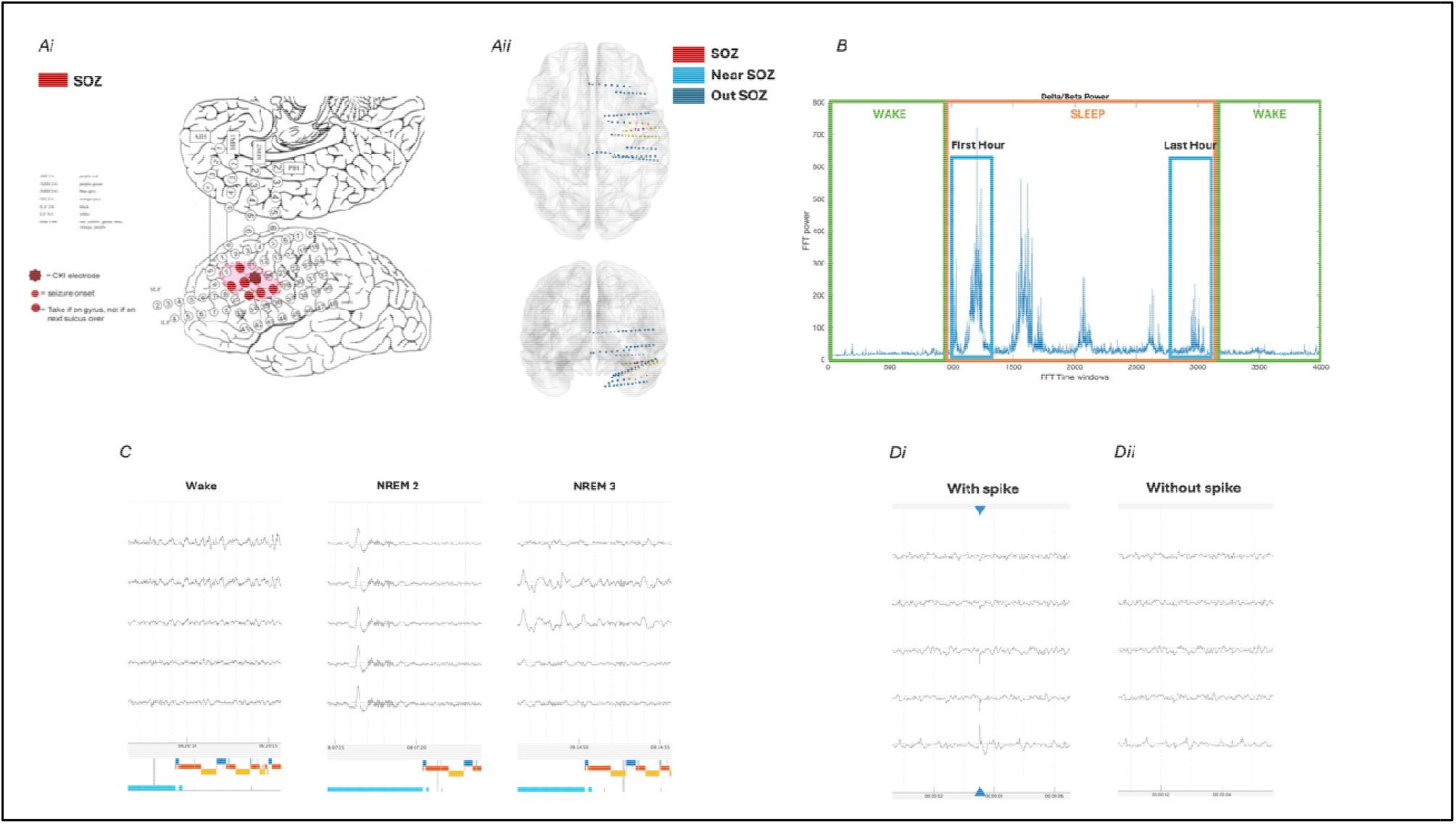
Summary of Methods. A. Patients included in this study consisted of well-characterized patients with who underwent intracranial recordings either with electrocorticogram (Ai) or with stereo-EEG electrodes (Aii). B. Sleep staging was based on a combination of whole-brain average delta-beta ratio plotting overnight and confirmation by visual sleep scoring of non-rapid eye movement (NREM) sleep epochs. The first and last hour of confirmed NREM sleep were then selected for data analysis. C. Representative intracranial EEG epochs of wakefulness, NREM stage 2 and NREM stage 3 sleep. D. Epileptiform discharges were visually identified and confirmed by a certified epileptologist and NREM sleep slow-wave activity was computed both with and without epileptic spikes. The slope of sleep slow waves vs. epileptic spike-waves were also separately analyzed and compared.

For this study, the SOZ electrodes for each patient were identified based on the clinically defined SOZ as well as based on the presence of gamma bursts 30 seconds before the seizures with the help of a qualified epileptologist (M.B.). Two centimeters of area around the SOZ electrodes was identified as “Near SOZ” area and the electrodes excluded from the “SOZ” and “Near SOZ” areas were identified as “Out SOZ”. For strip and grid electrodes Near SOZ electrodes consisted of electrodes within 2 cm radius of the SOZ electrodes with a representative patient in Figure 1Ai. For depth electrodes Near SOZ electrodes were identified to be lying within 2 cm Euclidean distance from the SOZ electrodes after electrode localization with a representative patient in Figure 1Aii.

### Data preprocessing

All the data processing and analysis was done using custom code on MATLAB (MathWorks, Natick, MA). For each patient, a whole night of recordings with no seizures at least six hours prior was selected. Nights were selected to have uninterrupted sleep cycles. These recordings were then manually sleep scored using the welch power in the delta (δ: 1-4 Hz) to beta (β: 12-25 Hz) ratio (Fig 1B) along with the presence of high amplitude slow-waves and spindles (Fig 1C). Interictal spikes were manually marked and verified by an epileptologist (M.B.) on each of these recordings (Fig 1Di). Bad channels and epochs were manually removed from all the recordings.

The recordings for each patient were divided to perform two analyses: 1) before removal of interictal spikes (with spikes) and 2) after removal of interictal spikes (without spikes) (Fig 1D). To analyze without spikes, 1-second epoch of data around the spikes was removed (Fig 1Dii).

The epochs of wake preceding and following sleep and the first hour and last hour of uninterrupted non-REM sleep cycles were identified from delta-beta ratio plots, then confirmed using visual sleep scoring (Fig 1B). If the uninterrupted sleep cycle was shorter than two hours in length, the data was divided into two equal parts for the analysis.

### Frequency band analysis

Welch power (6-second; non-overlapping windows) was calculated on both with and without spikes datasets to calculate the absolute δ power (slow wave activity [SWA]), beta power and the δ/β ratio overnight. Powers were then spatially z-scored across all electrodes. Absolute and relative δ decline was calculated using absolute δ powers as follows:

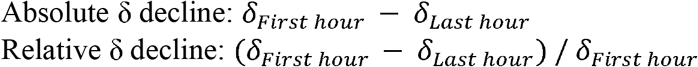

Average power values were calculated for both absolute and z-scored powers along with δ decline powers for electrodes in the SOZ, near SOZ and outside SOZ areas for further analysis. Please note that we will use delta power and SWA power interchangeably throughout the manuscript.

### Spike-wave and slow-wave slope analysis

Slow waves were marked by detecting negative peaks in each dataset after being placed through a bandpass filter between 0.5Hz and 4Hz. Any detected slow waves which occurred within 120ms following an interictal spike were considered spike waves. Slopes for each slow and spike waves were calculated before and after the zero crossing and were then averaged for each channel.

### Phase-Locked High Gamma (PLHG) calculation

Individual seizures were identified from each patient. Each patient had a varied number of seizures. These seizures were truncated to contain 30 seconds before the seizure onset to the end of the seizure and then analyzed. The PLHG measure was calculated based on a previous study by Weiss et.al. ^12,13^.

PLHG is the coupling between the phase in the lower frequencies 4 – 30 Hz and the amplitude of phase in the high gamma frequencies 80 – 150 Hz. For the PLHG calculations, the seizures were initially FIR filtered to the desired frequencies. Further, Hilbert transform was applied to calculate the respective phase and amplitudes and the values were normalized with respect to baseline (30 seconds before the seizure onset). PLHG was calculated for each 3 second time window with a 0.333 second overlap. PLHG was then averaged across each seizure for each individual patient.

### Statistical Analysis

Changes in absolute and z-scored SWA power for the SOZ, Near SOZ and Out SOZ were compared between wake and sleep within each region, then relative changes were directly compared across regions using paired t-tests. The z-scored SWA power and the relative change in δ and spike wave/slow wave slope in the SOZ, Near SOZ and Out SOZ were also compared for the first and the last hour of sleep for all the patients. A two-sample two tailed t-test was applied to compare the z-scored delta power and relative delta decline both with and without spikes for the different defined regions. The same test was applied to compare z-scored slopes of spike and slow waves as well as the changes in slope overnight.

To observe the differences in PLHG values between the SOZ, near SOZ and out SOZ a two sample two tailed t-test was applied. Average PLHG for each electrode from each patient was then Pearson correlated with the absolute δ power in the first hour, last hour and the relative δ decline for both with and without spikes. The average PLHG was similarly correlated with the slope of spike and slow waves in both the first and las hour as well as the relative slope change overnight. All statistics were thresholded for significance at false-discovery-rate (FDR) corrected p < 0.05 (with FDR estimated from p-values obtained across all comparisons described in the present work at once, all reported in Supplementary Material).

## Results

### SWA increases more from wake to sleep in the SOZ than in other areas

Absolute SWA power was quantified for each individual patient, with general framework displayed in a representative patient (P8) in Figure 2. Group-level analyses then quantified local differences in SWA during wake before sleep, the first and the last hours of sleep, wake after sleep, as well as local differences in overnight SWA decline in the cohort of sixteen patients with good quality intracranial EEG whole-night sleep data selected to be included in the present work (see Demographics in Table 1).

**Table 1:** Patient demographics.

| *Patient* | *Age Range* | *Gender* | *Electrode Type* | *Number of Electrodes Available* | *Epileptic Focus* |
| --- | --- | --- | --- | --- | --- |
| P1 | 26-30 | F | Depth electrodes | 59 | L. Mesial Temporal |
| P2 | 50-55 | F | Depth electrodes | 42 | R. Lateral Temporal |
| P3 | 20-25 | M | Depth electrodes | 23 | R. Mesial Temporal |
| P4 | 16-20 | M | Depth electrodes | 16 | R. Lateral Temporal |
| P5 | 20-25 | F | Depth electrodes | 24 | L. Lateral Temporal Pole (neocortical) |
| P6 | 20-25 | F | Depth electrodes | 16 | R. Lateral Temporal |
| P7 | 20-25 | F | Depth electrodes | 124 | L. Mesial Temporal |
| P8 | 20-25 | F | Depth electrodes | 96 | R. Lateral Temporal |
| P9 | 30-35 | M | Depth electrodes | 128 | R. Insula + Somatosensory Cortex |
| P10 | 50-55 | F | Depth electrodes | 128 | L. Mesial Temporal |
| P11 | 20-25 | M | Depth electrodes | 40 | R. Mesial Temporal |
| P12 | 30-35 | M | Depth electrodes | 16 | Bilateral Mesial Temporal |
| P13 | 36-30 | M | Strip and Grid Electrodes | 80 | L. Frontal Operculum |
| P14 | 20-25 | F | Strip and Grid Electrodes | 43 | L. Basal/Anterior Temporal |
| P15 | 16-20 | F | Strip and Grid Electrodes | 68 | R. Posterior Lateral Temporal |
| P16 | 26-30 | M | Strip and Grid Electrodes | 114 | L. Subtemporal/Lateral Temporal |

**Figure 2.**
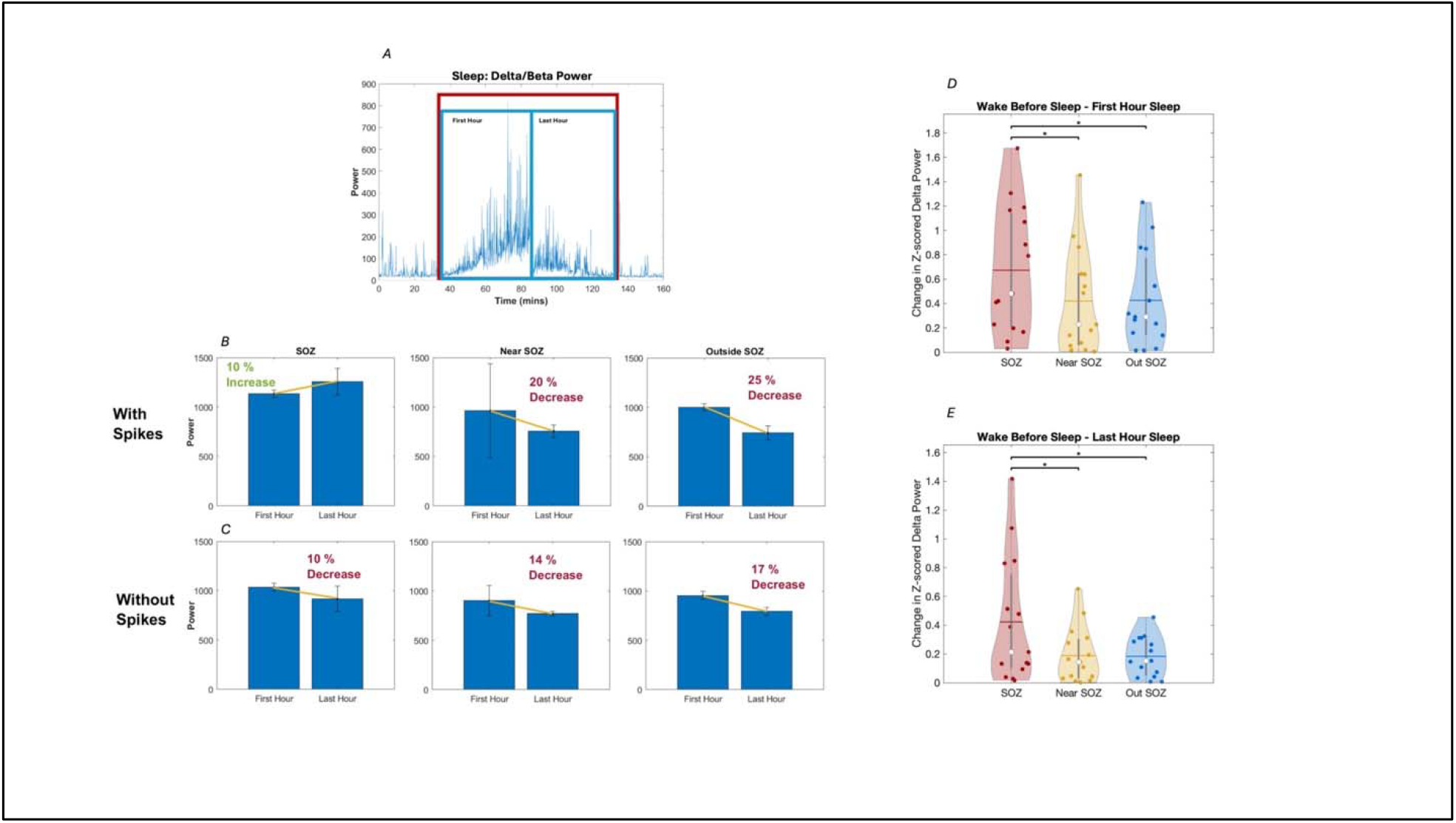
Illustration of sleep SWA analysis framework in a representative patient (P8), and group-level comparison of SWA levels between sleep and wake. (A) The first and last hour of uninterrupted sleep were selected based on average delta/beta power plots calculated across all regions. (B-C) Average SWA was then quantified separately for each region (in the SOZ, near the SOZ (<2 cm) and far from the SOZ (>2 cm)) for the first and last hour of sleep, and comparisons were made across regions both for SWA in early and late sleep and for overnight changes. Results were examined separately for data including vs excluding epileptic spikes. (D-E) Additional validation of sleep epoch selection was performed by comparing average sleep SWA between sleep and wake. These analyses showed that SWA increase from wake to sleep was higher in the SOZ compared to both neighboring regions (Near SOZ) and more distant regions (Out SOZ). Representative results here display changes in spatially z-scored SWA (delta power) between wake before sleep and the first and last hour of sleep, computed on data excluding epileptic spikes.

All regions (SOZ, Near SOZ, Out SOZ) showed significant increases in SWA during sleep compared to wakefulness (averaged across all epochs of sleep and wake). Absolute SWA changes were more consistent after removal of epileptic spikes from the dataset, while Z-scored changes were robustly observed in dataset containing or excluding spikes (Table S1).

Importantly, a direct comparison between regions showed consistently more increase in SWA from wakefulness to sleep in the SOZ compared to both neighboring areas (both for absolute and z-scored changes) and distant areas (for z-scored SWA changes). These changes were consistent whether including or removing epileptic spikes from the dataset (Table S2). Figure 1C-D, right panel, display results for Z-scored SWA changes between SOZ, Near SOZ and Out SOZ when comparing the first hour and the last hour of sleep to preceding wake,

### High local sleep SWA differentiates the SOZ from both neighboring and distant areas

Fig. 3 displays results of the group analysis comparing in z-scored SWA (emphasizing the consistency of relative topographical distribution) ^3^ calculated for the first hour of sleep, the last hour of sleep, as well as the relative change in SWA overnight. Z-scored SWA calculated when excluding epileptic spikes (Fig. 3A,B, Table S3) showed significantly higher power in the SOZ as compared to both regions neighboring it (FDR-corrected p = 0.0279 and 0.0215 for first and last hours of sleep respectively) and regions >2 cm radius from it (FDR-corrected p = 0.0279 and 0.0197 for first and last hours of sleep respectively). Similarly, Z-scored SWA calculated when including epileptic spikes (Fig. 3C,D, Table S3) showed significantly higher power in the SOZ as compared to regions neighboring it (FDR-corrected p = 0.0215 and 0.0197 for the first and last hours of sleep respectively) and regions >2 cm away from it FDR-corrected p = 0.0215 and 0.0197 for the first and last hours of sleep). Group analysis on z-scored slopes of sleep slow waves also revealed greater slope magnitude in the SOZ compared to both areas near it and far from it (Fig. 3E,F, Table S3). Similar results were obtained for epileptic spike-waves (Fig. 3 G,H, Table S3). Unlike for SWA power, there was however no significant difference in sleep slow-wave slope or epileptic spike-wave slope between the areas neighboring the SOZ and the areas far from it. Overall similar results were observed when using absolute SWA values rather than Z-scored values (not adjusting for inter-subject average value differences) when comparing both SWA power and slow-wave slopes during the first and the last hours of sleep, but results did not reach significance as consistently (Supplementary Fig. 1, Table S3).

**Figure 3.**
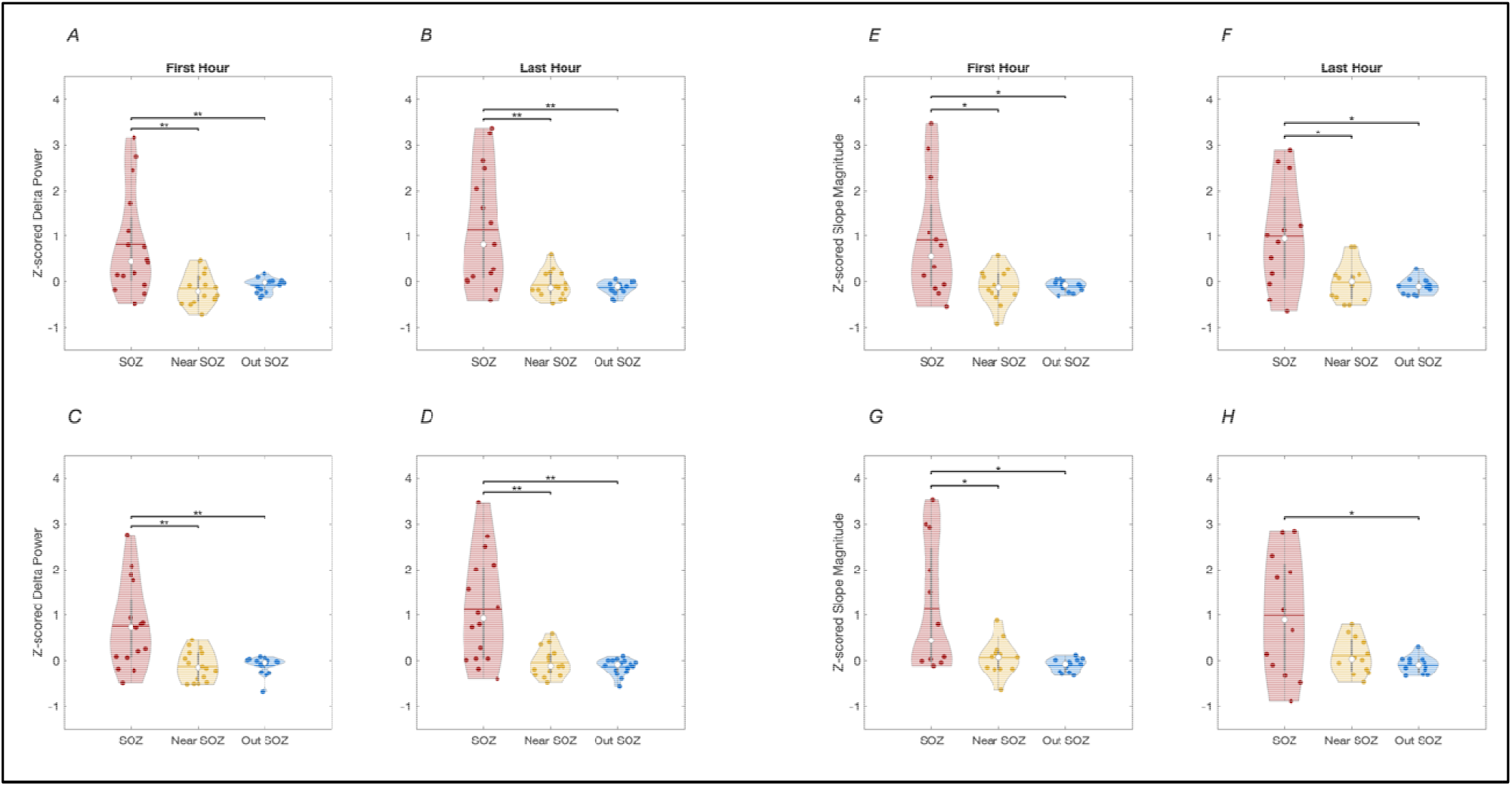
Group results for regional differences in SWA (i.e. Delta Power, 1-4 Hz) and slope of sleep slow-waves vs epileptic spike-waves across the seizure onset zone (SOZ, red), brain areas less than two centimeters away (near SOZ, yellow) and brain areas more than two centimeters away (Out SOZ, blue). Spatially Z-scored are displayed, highlighting consistent differences in topography while minimizing effects of inter-individual differences in absolute power. A and B: SWA (Delta Power) during the first and last hours of NREM sleep, data excluding epileptic spikes. C and D: SWA (Delta Power) during the first and last hours of NREM sleep, data including epileptic spikes. E and F: Sleep slow-wave slope during the first and last hours of NREM sleep, data excluding epileptic spikes. G and H: Epileptic spike-wave slope during the first and last hours of NREM sleep. ** denote p<0.01 and * p<0.05 (FDR-corrected).

### Overnight decline in sleep SWA is also dampened in the SOZ

Before examining regional differences in overnight decline in sleep SWA, we first validated that sleep SWA was declining overall in the whole brain, as has been consistently observed during physiological sleep in both animals and human studies. At the group level, across all sixteen patients, there was a indeed significant overall decline in SWA overnight (Table S4), with the highest decline far from the SOZ (36% decline, p = 0.0056), relatively lower decline near the SOZ (27% decline, p = 0.0215), and the least decline or even slight increase in SWA overnight in some patients in the SOZ (27% decline, p = 0.011). An overnight decrease in slope magnitude for sleep slow waves was also observed in all areas, although the slope of slow waves outside the SOZ trended towards greater change averaging a 17% decrease (p = 0.0226). The slope of slow waves near and within the SOZ also displayed a 12% decrease but did not reach significance. In contrast to sleep slow-wave slopes, the slopes of epileptic spike-wave did not display an overall decrease overnight (Table S4).

Group analysis of relative overnight decline in SWA (normalized for each region by absolute power values for the first and last hour of sleep) showed a lower decline in the SOZ as compared to far from the SOZ (p = 0.0303) (Fig. 4, Table S5). No significant difference was observed between the SOZ and neighboring areas. Although a similar pattern was observed, no significant differences were observed in overnight decline of slope of slow-wave or spike-waves between any of the regions of interest (Fig 4, Table S5).

**Figure 4.**
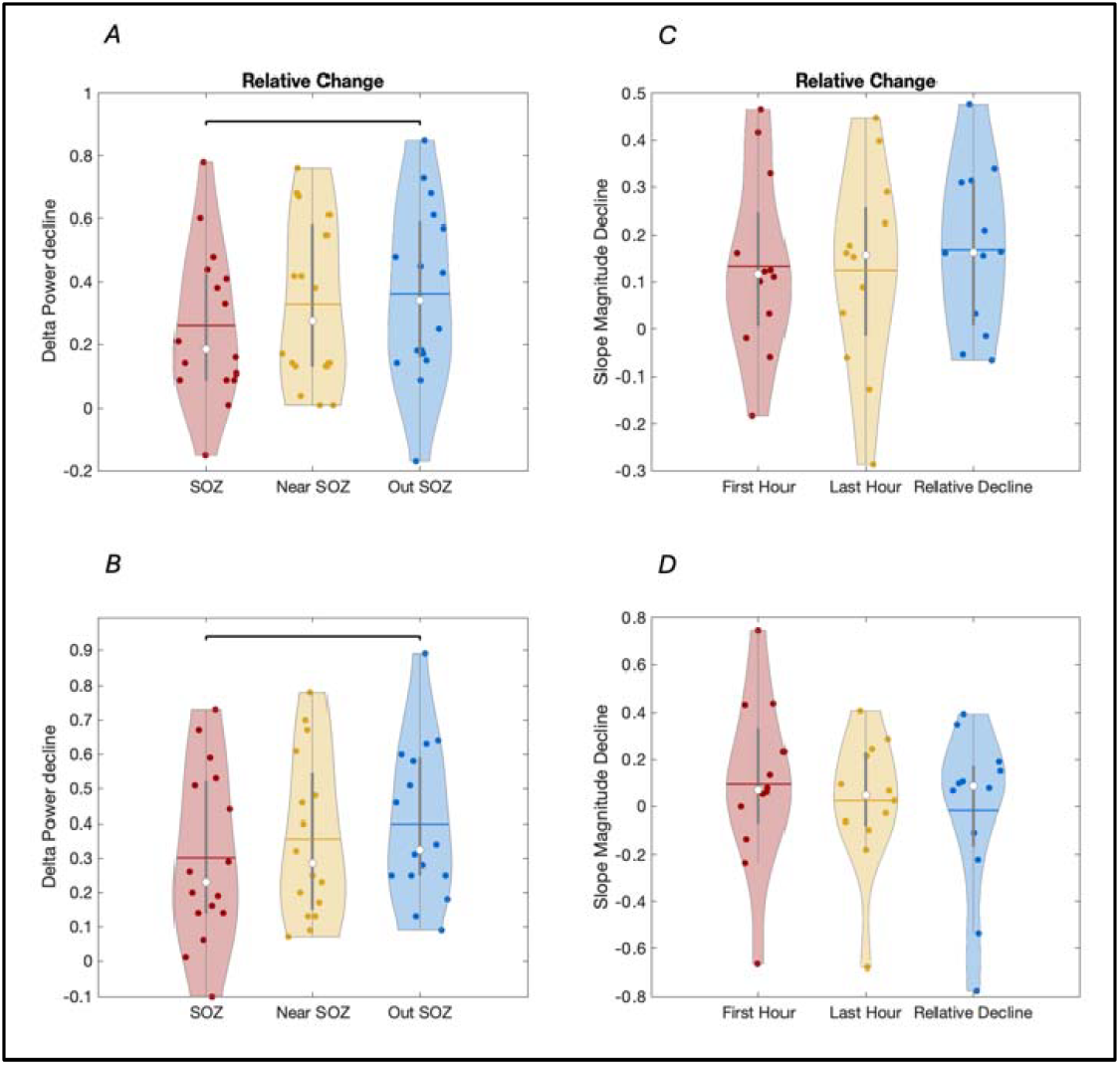
Group results for regional differences in overnight decline in SWA (i.e. Delta Power, 1-4 Hz) and in the slope of sleep slow-waves vs epileptic spike-waves across the seizure onset zone (SOZ, red), brain areas less tha two centimeters away (near SOZ, yellow) and brain areas more than two centimeters away (Out SOZ, blue). Absolute power differences in power are here displayed. A: Overnight decline in SWA (Delta Power), data excluding epileptic spikes. B: Overnight decline in SWA (Delta Power), data including epileptic spikes. C: Overnight decline in sleep slow-wave slope, data excluding epileptic spikes. G and H: Overnight decline in epileptic spike-wave slope. * denote p<0.05 (FDR-corrected).

### Relationship of PLHG with SWA, sleep slow-wave and spike-wave slopes

Fig. 5a displays the time course of PLHG values across all channels during a seizure for a representative patient (P15). As a methodological validation for PLHG, group-level statistics (Fig. 5b) also showed that PLHG was significantly higher in the SOZ as compared to areas neighboring to it (p < 0.001) and far from it (p < 0.01).

**Figure 5.**
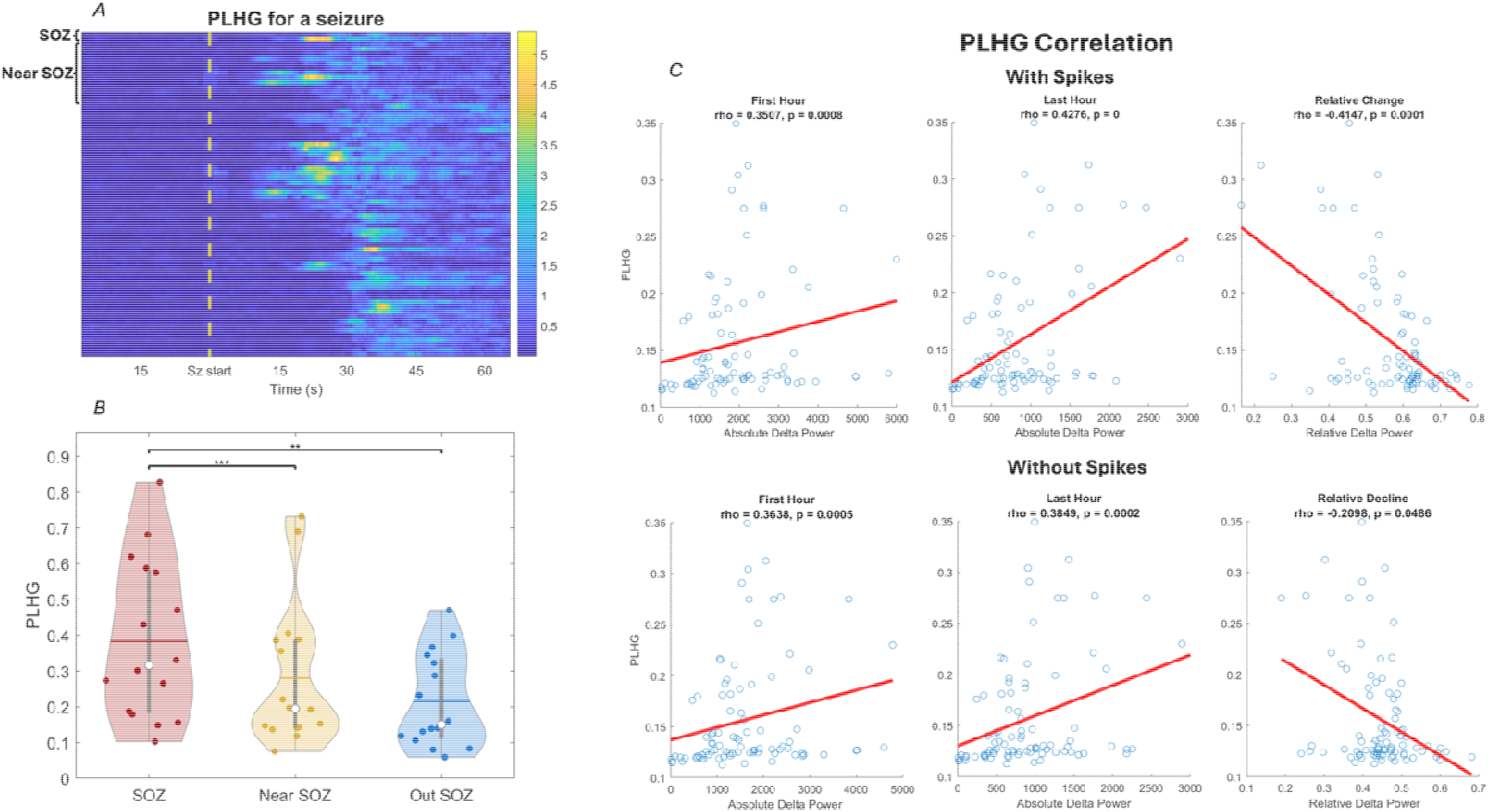
PLHG analysis framework and example of individual results in representative subject results. One can already visually identify an overall higher PLHG within the SOZ and near the SOZ as compared to the rest of the brain during the whole seizure. One can see that PLHG is positively correlated with the absolute SWA power (data including spikes) in the first hour (r = 0.3507, p < 0.001) and the last hour (r = 0.4276, p < 0.001), and negatively correlated with the relative decline in SWA overnight (r = -0.4141, p < 0.001) (Fig 4c i-iii). Results were similar for absolute SWA power calculated from data without spikes: PLHG positively correlated with the SWA power in the first hour (r = 0.3638, p = 0.0005) and the last hour (r = 0.3849, p = 0.0002) of sleep, and negatively correlated with the relative decline in SWA overnight (r = -0.2098, p = 0.048). Similar results were also observed for the correlation of PLHG with Z-scored SWA in the first hour (with spikes: r = 0.3507, p = 0.0008; without spikes: r = 0.3638, p = 0.0004) and last hour (with spikes: r = 0.4275, p = 0.00003; without spikes: r = 0.3849, p = 0.0002), with spikes as well as without spikes. PLHG also displayed a comparable positive correlation with the slope of both spike and slow waves during the first and last hours of sleep, while the relative change in slope magnitude overnight displayed weak positive correlation which did not reach significance in this single-subject analysis.

To illustrate our correlation approach, single-subject results for an association between PLHG and SWA power across channels is also plotted in representative patient P15 in Fig. 5c. Group level results (Fig. 6, Table S6) revealed a positive trending correlation between ictal PLHG and local SWA power in sleep data both without inclusion of epileptic spikes (trending in the first hour (uncorrected p = 0.0353) and significant in the last hour (FDR-corrected p = 0.0279)) and when including them (trending in the first hour (uncorrected p = 0.0807) and significant in the last hour (FDR-corrected p = 0.0315)). Ictal PLHG also displayed a positive correlation with the slope of both spike-waves (in the first (p = 0.0303) and the last hour (p = 0.0295)) and sleep slow-waves (in the first (p = 0.0239) and the last hour (p = 0.0215)) at the group level. Finally, ictal PLHG was also trended negatively with the relative decline in SWA power over the course of the night. However, the correlation between ictal PLHG and the overnight change in slope of spike-waves or sleep-slow waves did not reach significance after FDR correction (Table S6).

**Figure 6.**
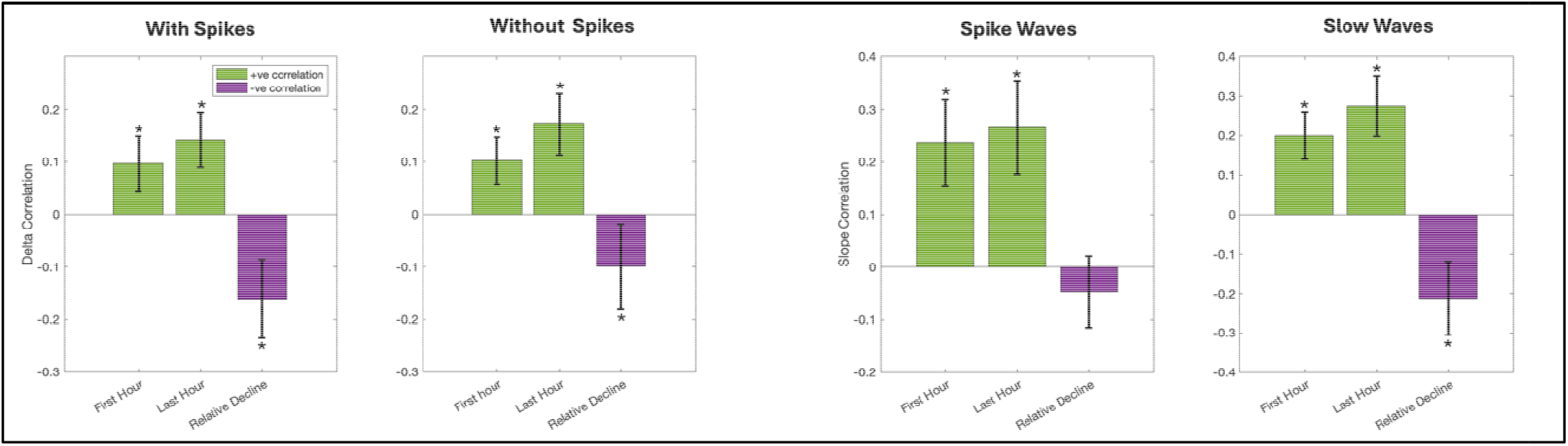
Group results for correlation analysis between intracranial EEG markers of sleep homeostasis (sleep SWA, slope of sleep slow-waves and epileptic spike-waves) and ictal PLHG (a marker of local ictal recruitment). A. Correlation between sleep SWA (i.e. Delta Power) and ictal PLHG, data without epileptic spikes. B. Correlatio between sleep SWA (i.e. Delta Power) and ictal PLHG, data with epileptic spikes. C. Correlation between the slop of sleep slow-waves and ictal PLHG, data with epileptic spikes. D. Correlation between the slope of epileptic spike-waves and ictal PLHG. Y axis displays correlation values. Group mean and standard error are plotted for eac analysis. * denotes p<0.05 (FRD-corrected).

## Discussion

Using direct intracranial EEG recordings from sixteen well-characterized human patients with focal epilepsy (FE), we showed that locally increased sleep slow-wave activity (SWA) is a promising interictal biomarker for the seizure onset zone (SOZ). SWA in the SOZ increased more from wake to sleep than in both neighboring or distant regions. Within sleep, locally increased SWA also reliably differentiated the SOZ from neighboring regions, especially during the last hour of nighttime sleep. Importantly, increases in sleep SWA in the SOZ were also found to be associated with a dampened overnight decline in local sleep SWA, suggesting that the epileptic focus escapes from physiological sleep homeostasis. Across all regions, local abnormalities in markers of sleep homeostasis were found to be directly proportional to ictal recruitment, as measured by phase-locked high-gamma (PHLG) activity. Overall, these results support the hypothesis that local disruptions in sleep homeostasis may contribute to the persistence of local hyperexcitability in epileptic brains from night to night, and thus to recurrence of refractory seizures often observed in patients with FE.

The present results show that SWA increases more from wakefulness to sleep in the SOZ compared to both neighboring and distant regions. These findings are consistent with—and extend—our previous scalp EEG results showing greater SWA increases in non-REM sleep compared to REM sleep in patients with FE, here demonstrating that this effect is especially pronounced within the SOZ itself.^29^ This supports the hypothesis that the mechanisms underlying sleep-enhanced focal ‘slowing’ in epileptic networks may differ from the typical disconnection phenomena observed, for example, in stroke patients, and may instead reflect a homeostatic response to local increases in synaptic strength, high-jacking normal plasticity mechanisms.^29^ This interpretation aligns with human slice data showing a 2.6-fold increase in synaptic density, accompanied by elevated CREB expression (a marker of long-term potentiation), in layer 2/3 of epileptic neocortex relative to adjacent cortex. ^9^ Future work may directly relate local increases in intracranial sleep SWA to postoperative histological measures of synaptic density in the same subjects.

Our results also suggest that sleep SWA remained increased throughout the night in the SOZ compared to neighboring and distant areas, with the most consistent results observed towards the end of the night. The presence of locally increased sleep SWA aligns with previous evidence of increased synaptic density, especially marked in the superficial layers, in the SOZ in human tissue samples.^9-11^ The observed persistence of increased sleep SWA at the end of the night contrasts with previous findings of normal increases in sleep SWA following physiological learning, which rapidly returns to baseline within the first hour of sleep. ^28^ In contrast, our results also show that overnight sleep SWA decline is dampened in the SOZ compared to the rest of the brain. Given the propensity of physiological synaptic pruning to mostly target smaller synapses,^18^ persistent increases in sleep SWA in epileptic networks may result from the presence of strongly potentiated synapses in the epileptic focus, effectively hijacking typically adaptive mechanisms. ^29^ Frequent interictal epileptiform discharges (IEDs) during sleep, which are typically more abundant near the SOZ and/or at the SOZ itself compared to the rest of the brain, may also contribute to this phenomenon. ^29^ Indeed, animal studies suggest that burst firing patterns (as found during epileptic spikes ^14^) may prevent synaptic pruning related to slow-wave activity, with long-lasting preventive effects observed ^22^. A preventive effect of bursting during spikes on synaptic pruning is also supported by our finding of an absence of overnight decline in the slopes of epileptic spike-waves (as opposed to those of sleep slow waves). Overall, the present results suggest a local dysfunction in sleep homeostatic processes in the epileptic brain, maximal in the SOZ, that cannot be simply accounted for by global sleep disturbances such as sleep apnea or insomnia in these patients. These results strongly suggest the potential relevance of local neuromodulation approaches targeting the SOZ to enhance local sleep homeostasis (e.g., through low-frequency auditory stimulation ^30^ or deep brain stimulation ^31^ during sleep).

Our present findings of a direct correlation between increases in sleep SWA and slow-wave slope and local ictal recruitment as measured by ictal PHLG are in line with our previous, more indirect scalp-level results of an association between whole-scalp sleep SWA and the frequency of seizures with impaired awareness captured in the same EMU stay. ^29^ Such a selective association between widespread increases in sleep SWA and impaired awareness during seizures also aligns with our recent findings of higher diffuse increases in ictal PHLG during focal seizures with secondary generalization compared to seizures with impaired awareness that remain focal. ^32^ Overall, these results suggest that focal seizures, especially those leading to impaired awareness, may lead to widespread maladaptive increases in synaptic strength in the human brain that may compete with normal learning mechanisms and secondarily worsen sleep homeostasis, further contributing to seizure refractoriness in a closed-loop deleterious cycle. Future studies may compare strategies to enhance sleep homeostasis at the level of the whole brain (e.g., using auditory stimulation or targeting core sleep regulation centers such as the thalamus; ref) versus at the local level of the epileptogenic network in terms of improvement in both seizure frequency and cognitive outcomes.

## Limitations

The current study does have several limitations. Intracranial recordings cannot span the whole cortex, and there was no subcortical coverage of arousal centers such as the thalamus in the present sample. Sleep staging was performed without a full polysomnography at the scalp level, and as a result, REM sleep was unable to be studied separately. The patient cohort was not large enough to allow for powered analyses to detect differences between SOZs in different cortical lobes. Finally, larger studies should also assess whether decreased overnight SWA decline in the SOZ predicts surgical outcomes.

## Conclusion

Our results confirm our previous scalp-level findings of focal impairment of sleep homeostasis in patients with FE, with direct evidence for a focal impairment at the SOZ in human patients with intracranial EEG data. Future studies may seek to enhance sleep homeostasis in the SOZ via direct cortical stimulation or through non-invasive targeted neuromodulation, such as transcranial electrical stimulation with temporal interference, to further causally relate improvements in sleep homeostasis with modified clinical outcomes.

## Data Availability

All data produced in the present study are available upon reasonable request to the authors

## Acknowledgments

This work was supported by NINDS K23NS112473. G.T. holds an executive position in Intrinsic Powers, Inc., a company whose purpose is to develop a device that can be used in the clinic to assess the presence and absence of consciousness in patients. Vaclav Kremen has a consulting relationship with Certicon and has intellectual property related to behavioral state and seizure classification algorithms.

