## Supplementary Material for "Direct intracranial EEG evidence for a local breakdown in normal sleep homeostasis at the human seizure onset zone"

**Table S1: Within-region Comparisons of Sleep and Wake SWA**

***Z-scored values***

Table S1a:

|  | *Z-Scored SWA Power Without Spikes: SOZ* | | |
| --- | --- | --- | --- |
|  | | *Wake* | *Sleep* |
| Mean | | 0.102089517 | 0.483582169 |
| SEM | | 0.115822785 | 0.097257932 |
| P-Value | | **0.0201** | |

Table S1b:

|  | *Z-Scored SWA Power Without Spikes: Near SOZ* | | |
| --- | --- | --- | --- |
|  | | *Wake* | *Sleep* |
| Mean | | -0.156409063 | 0.120549219 |
| SEM | | 0.054595083 | 0.059092993 |
| P-Value | | **0.0074** | |

Table S1c:

|  | *Z-Scored SWA Power Without Spikes: Outside SOZ* | | |
| --- | --- | --- | --- |
|  | | *Wake* | *Sleep* |
| Mean | | -0.182286033 | 0.126698656 |
| SEM | | 0.041679555 | 0.031052493 |
| P-Value | | **0.0063** | |

Table S1d:

|  | *Z-Scored SWA Power With Spikes: SOZ* | | |
| --- | --- | --- | --- |
|  | | *Wake* | *Sleep* |
| Mean | | 0.308389553 | 0.851787625 |
| SEM | | 0.167794891 | 0.185211269 |
| P-Value | | **0.0199** | |

Table S1e:

|  | *Z-Scored SWA Power With Spikes: Near SOZ* | | |
| --- | --- | --- | --- |
|  | | *Wake* | *Sleep* |
| Mean | | -0.185450667 | 0.125047094 |
| SEM | | 0.050196861 | 0.059604593 |
| P-Value | | **0.0032** | |

Table S1f:

|  | *Z-Scored SWA Power With Spikes: Outside SOZ* | | |
| --- | --- | --- | --- |
|  | | *Wake* | *Sleep* |
| Mean | | -0.2179817 | 0.103224113 |
| SEM | | 0.03378592 | 0.027908984 |
| P-Value | | **0.0013** | |

***Absolute values***

Table S1g:

|  | *SWA Power Without Spikes: SOZ* | | |
| --- | --- | --- | --- |
|  | | *Wake* | *Sleep* |
| Mean | | 892.2893333 | 1445.24 |
| SEM | | 214.4780399 | 297.7031627 |
| P-Value | | **0.0118** | |

Table S1h:

|  | *SWA Power Without Spikes: Near SOZ* | | |
| --- | --- | --- | --- |
|  | | *Wake* | *Sleep* |
| Mean | | 530.0159 | 817.3271875 |
| SEM | | 186.4025931 | 243.5168174 |
| P-Value | | **0.0185** | |

Table S1i:

|  | *SWA Power Without Spikes: Outside SOZ* | | |
| --- | --- | --- | --- |
|  | | *Wake* | *Sleep* |
| Mean | | 620.0334667 | 1019.080313 |
| SEM | | 241.9487318 | 365.4490203 |
| P-Value | | **0.0285** | |

Table S1j:

|  | *SWA Power With Spikes: SOZ* | | |
| --- | --- | --- | --- |
|  | | *Wake* | *Sleep* |
| Mean | | 2698.657333 | 3500.430313 |
| SEM | | 1815.493759 | 2087.434577 |
| P-Value | | **0.0397** | |

Table S1k:

|  | *SWA Power With Spikes: Near SOZ* | | |
| --- | --- | --- | --- |
|  | | *Wake* | *Sleep* |
| Mean | | 2225.907 | 2779.942188 |
| SEM | | 1878.305323 | 2176.271172 |
| P-Value | | 0.1003 | |

Table S1l:

|  | *SWA Power With Spikes: Outside SOZ* | | |
| --- | --- | --- | --- |
|  | | *Wake* | *Sleep* |
| Mean | | 2324.7127 | 3002.046031 |
| SEM | | 1963.317088 | 2321.76802 |
| P-Value | | 0.0978 | |

**Table S2: Regional Differences in SWA Change between Sleep and Wake**

Table S2a:

| Change in SWA Power: Without Spikes | Wake Before Sleep > First Hour of Sleep | Wake Before Sleep > Last Hour of Sleep | Wake After Sleep > First Hour of Sleep | Wake After Sleep > Last Hour of Sleep |
| --- | --- | --- | --- | --- |
| SOZ vs Near SOZ | **0.027** | **0.03** | 0.061 | **0.0363** |
| SOZ vs Outside SOZ | 0.1711 | 0.0965 | 0.171 | 0.1916 |
| Near SOZ vs Outside SOZ | 0.1756 | 0.0954 | 0.1308 | 0.1078 |

Table S2b:

| Change in Z-Scored SWA Power: Without Spikes | Wake Before Sleep > First Hour of Sleep | Wake Before Sleep > Last Hour of Sleep | Wake After Sleep > First Hour of Sleep | Wake After Sleep > Last Hour of Sleep |
| --- | --- | --- | --- | --- |
| SOZ vs Near SOZ | **0.0315** | **0.0421** | **0.0478** | 0.0602 |
| SOZ vs Outside SOZ | **0.0397** | **0.0402** | 0.0987 | 0.099 |
| Near SOZ vs Outside SOZ | 0.4262 | 0.4192 | 0.2011 | 0.076 |

Table S2c:

| Change in SWA Power: With Spikes | Wake Before Sleep > First Hour of Sleep | Wake Before Sleep > Last Hour of Sleep | Wake After Sleep > First Hour of Sleep | Wake After Sleep > Last Hour of Sleep |
| --- | --- | --- | --- | --- |
| SOZ vs Near SOZ | **0.0368** | **0.0296** | 0.0837 | 0.0539 |
| SOZ vs Outside SOZ | 0.0972 | 0.0964 | 0.1711 | 0.2587 |
| Near SOZ vs Outside SOZ | 0.1582 | 0.195 | 0.1063 | 0.1239 |

Table S2d:

| Change in Z-Scored SWA Power: With Spikes | Wake Before Sleep > First Hour of Sleep | Wake Before Sleep > Last Hour of Sleep | Wake After Sleep > First Hour of Sleep | Wake After Sleep > Last Hour of Sleep |
| --- | --- | --- | --- | --- |
| SOZ vs Near SOZ | **0.0257** | **0.0318** | **0.0304** | **0.0298** |
| SOZ vs Outside SOZ | **0.0262** | **0.03** | **0.0326** | **0.0322** |
| Near SOZ vs Outside SOZ | 0.3453 | 0.2489 | 0.4113 | 0.3146 |

**Table S3: Regional differences in sleep SWA, slow-wave and spike-wave slopes**

***Z-scored values***

Table S3a:

| Z-Scored SWA: Without Spikes | First Hour | Last Hour |
| --- | --- | --- |
| SOZ vs Near SOZ | **0.0245** | **0.0178** |
| SOZ vs Outside SOZ | **0.0240** | **0.0123** |
| Near SOZ vs Outside SOZ | 0.3095 | 0.2639 |

Table S3b:

| Z-Scored SWA: With Spikes | First Hour | Last Hour |
| --- | --- | --- |
| SOZ vs Near SOZ | **0.0179** | **0.0126** |
| SOZ vs Outside SOZ | **0.0178** | **0.0126** |
| Near SOZ vs Outside SOZ | 0.4293 | 0.1919 |

Table S3c:

| Z-Scored Slope Amplitude: Slow Waves | First Hour | Last Hour |
| --- | --- | --- |
| SOZ vs Near SOZ | **0.0299** | **0.0262** |
| SOZ vs Outside SOZ | **0.0279** | **0.0174** |
| Near SOZ vs Outside SOZ | 0.4420 | 0.3103 |

Table S3d:

| Z-Scored Slope Amplitude: Spike Waves | First Hour | Last Hour |
| --- | --- | --- |
| SOZ vs Near SOZ | **0.0299** | 0.0433 |
| SOZ vs Outside SOZ | **0.0198** | **0.0253** |
| Near SOZ vs Outside SOZ | 0.1311 | 0.1002 |

***Absolute values***

Table S3e:

| SWA Absolute Values: Without Spikes | First Hour | Last Hour |
| --- | --- | --- |
| SOZ vs Near SOZ | 0.4303 | 0.4236 |
| SOZ vs Outside SOZ | 0.3429 | 0.3414 |
| Near SOZ vs Outside SOZ | 0.4260 | 0.1899 |

Table S3f:

| SWA Absolute Values: With Spikes | First Hour | Last Hour |
| --- | --- | --- |
| SOZ vs Near SOZ | 0.4291 | 0.4284 |
| SOZ vs Outside SOZ | 0.3271 | 0.3434 |
| Near SOZ vs Outside SOZ | 0.3105 | 0.3115 |

Table S3g:

| Slope Amplitude Absolute Values: Slow Waves | First Hour | Last Hour |
| --- | --- | --- |
| SOZ vs Near SOZ | 0.3670 | 0.3815 |
| SOZ vs Outside SOZ | 0.3734 | 0.3801 |
| Near SOZ vs Outside SOZ | 0.4414 | 0.4437 |

Table S3h:

| Slope Amplitude Absolute Values: Slow Waves | First Hour | Last Hour |
| --- | --- | --- |
| SOZ vs Near SOZ | 0.2006 | 0.3059 |
| SOZ vs Outside SOZ | 0.1861 | 0.2762 |
| Near SOZ vs Outside SOZ | 0.4260 | 0.4248 |

**Table S4: overnight decline in SWA, slow-wave slope and spike-wave slope**

Table S4a: SWA

|  | *SOZ* | *SOZ 2cm* | *Out SOZ* |
| --- | --- | --- | --- |
| Mean | -27% | -29% | -36% |
| SEM | 5.967075 | 8.279225 | 6.990004 |
| P-Value | **0.0056** | **0.0187** | **0.0024** |

Table S4b: slow-wave slope

|  | *SOZ* | *SOZ 2cm* | *Out SOZ* |
| --- | --- | --- | --- |
| Mean | -12% | -12% | -17% |
| SEM | 0.05515 | 0.06138 | 0.04959 |
| P-Value | 0.0403 | 0.0638 | **0.0207** |

Table S4c: spike-wave slope

|  | *SOZ* | *SOZ 2cm* | *Out SOZ* |
| --- | --- | --- | --- |
| Mean | -10% | -3% | 2% |
| SEM | 0.10408 | 0.08122 | 0.0999 |
| P-Value | 0.2418 | 0.3866 | 0.4274 |

**Table S5: Regional differences in overnight decline**

| Overnight Relative Decline | SWA: without spikes | SWA: with spikes | Slow-wave slope | Spike-wave slope |
| --- | --- | --- | --- | --- |
| SOZ vs Near SOZ | 0.0599 | 0.0987 | 0.4251 | 0.3310 |
| SOZ vs Outside SOZ | **0.0206** | **0.0255** | 0.3389 | 0.2721 |
| Near SOZ vs Outside SOZ | 0.1645 | 0.0636 | 0.3284 | 0.3806 |

**Table S6: Correlation with PLHG**

Table S6a:

|  | *PLHG Correlation: SWA Without Spikes* | | | *PLHG Correlation: SWA With Spikes* | | |
| --- | --- | --- | --- | --- | --- | --- |
|  | *First Hour* | *Last Hour* | *Relative Decline* | *First Hour* | *Last Hour* | *Relative Decline* |
| T-Values | 2.3138 | 2.9766 | -1.2181 | 1.8728 | 2.7153 | -2.1479 |
| SEM | 0.0444 | 0.0578 | 0.0814 | 0.0518 | 0.0523 | 0.0748 |
| P-Value | **0.0405** | **0.0247** | 0.1715 | 0.0760 | **0.0280** | 0.0510 |

Table S6b:

|  | *PLHG Correlation: Slow Waves Slope Amplitude* | | | *PLHG Correlation: Spike Waves Slope Amplitude* | | |
| --- | --- | --- | --- | --- | --- | --- |
|  | *First Hour* | *Last Hour* | *Relative Decline* | *First Hour* | *Last Hour* | *Relative Decline* |
| T-Values | 3.324 | 3.5808 | -2.2933 | 2.8936 | 2.9784 | -0.6854 |
| SEM | 0.0603 | 0.0768 | 0.0927 | 0.0819 | 0.0893 | 0.0680 |
| P-Value | **0.0195** | **0.0181** | **0.0462** | **0.0271** | **0.0265** | 0.3047 |

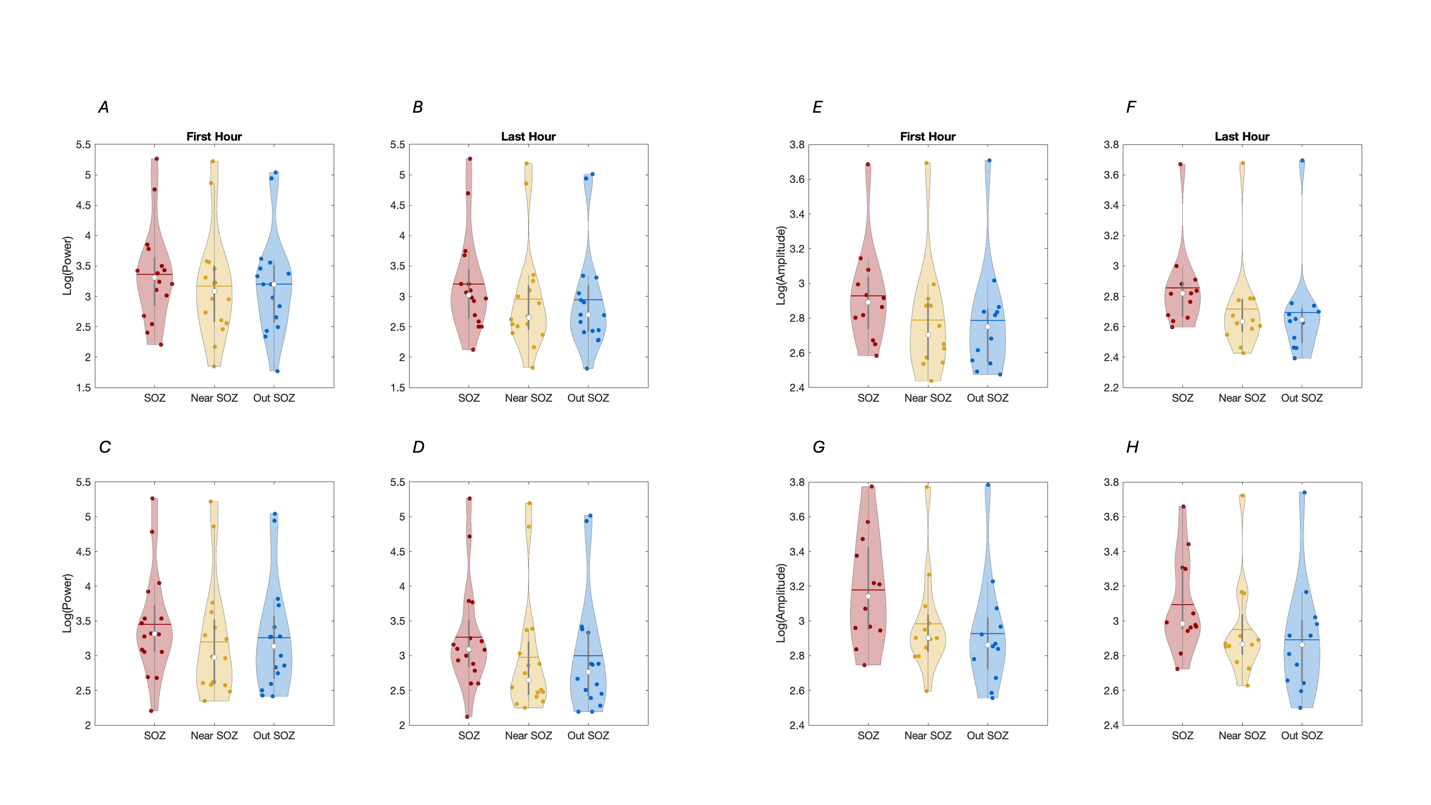
**Supplementary Figure 1:** Group results for regional differences in SWA (i.e. Delta Power, 1-4 Hz) and slope of sleep slow-waves vs epileptic spike-waves across the seizure onset zone (SOZ, red), brain areas less than two centimeters away (near SOZ, yellow) and brain areas more than two centimeters away (Out SOZ, blue). Absolute values are displayed. A and B: SWA (Delta Power) during the first and last hours of NREM sleep, data excluding epileptic spikes. C and D: SWA (Delta Power) during the first and last hours of NREM sleep, data including epileptic spikes. E and F: Sleep slow-wave slope during the first and last hours of NREM sleep, data excluding epileptic spikes. G and H: Epileptic spike-wave slope during the first and last hours of NREM sleep. While similar trends were observed using absolute values as opposed to Z-scored data (as in main Figure 3), no significant results were here obtained at the group level.

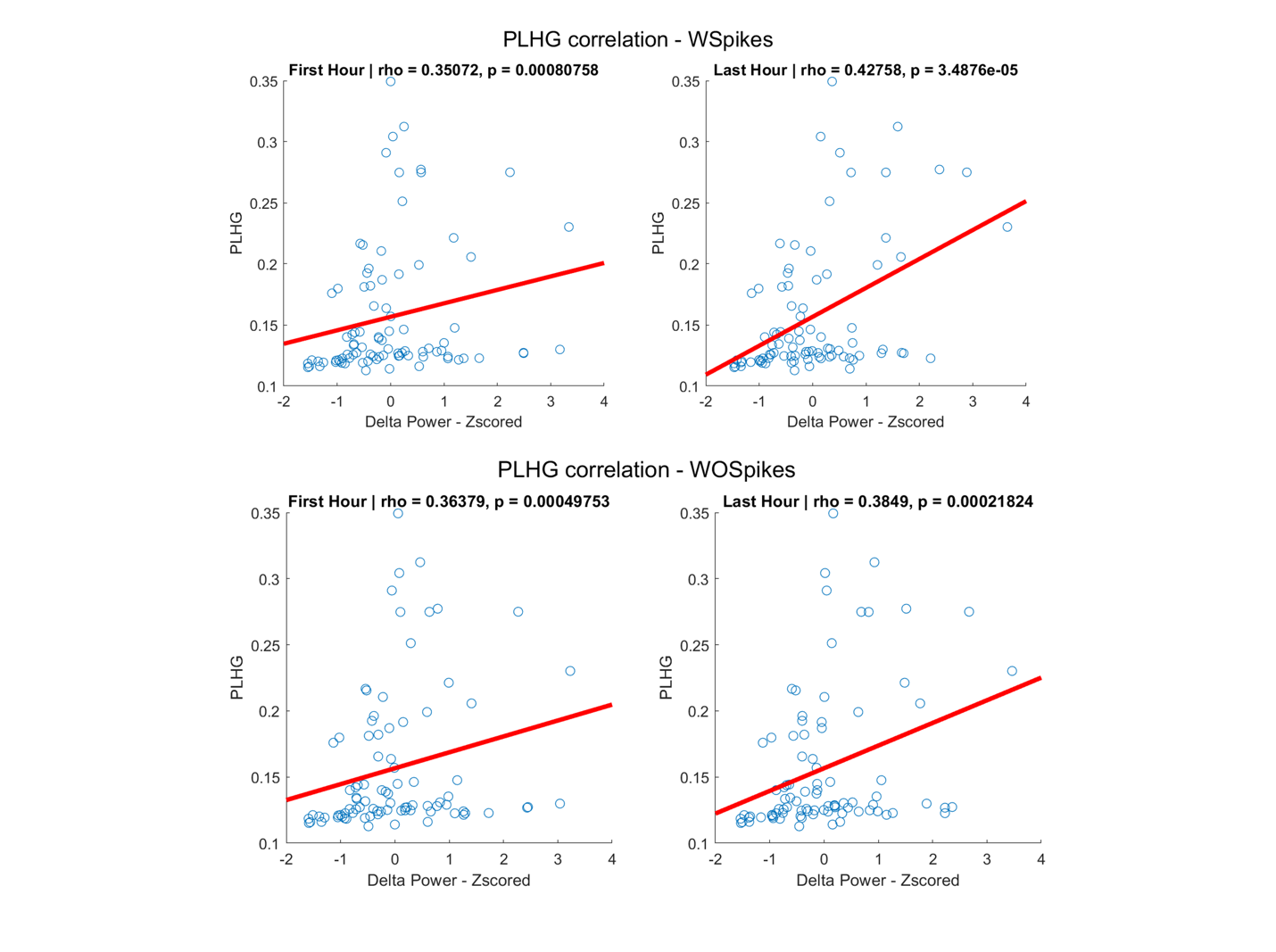

**Supplementary Figure 2:** results from a representative subject for correlation between PLHG and z-scored delta power (SWA).
